# Cohort Profile: The Australian Generating evidence on antimicrobial resistance in the aged care environment (GRACE) study; alignment with national population characteristics

**DOI:** 10.1101/2022.10.26.22281199

**Authors:** Lucy Carpenter, Andrew Shoubridge, Erin Flynn, Catherine Lang, Steven Taylor, Lito Papanicolas, Josephine Collins, David Gordon, David J. Lynn, Maria Crotty, Craig Whitehead, Lex Leong, Steve Wesselingh, Kerry Ivey, Maria C. Inacio, Geraint Rogers

**Author notes:** **Corresponding author:** Lucy Carpenter, Microbiome and Host Health Programme, 5D332 Flinders Medical Centre, University Drive, Bedford Park SA 5042 Australia. These authors contributed equally to this work (Maria C. Inacio, Geraint Rogers).

## Abstract

**Purpose:** The emergence of antibiotic-resistant bacteria represents a considerable threat to human health, particularly for vulnerable populations such as those living in residential aged care. However, antimicrobial resistance (AMR) carriage and modes of transmission remain incompletely understood. The Generating evidence on antimicrobial Resistance in the Aged Care Environment (GRACE) study was established to determine principal risk factors of AMR carriage and transmission in residential aged care facilities (RACF).

**Participants:** Between March 2019 and March 2020, 279 participants were recruited from five South Australian RACFs. The median age was 88.6 years, the median period in residence was 681 days, and 71.7% were female. A dementia diagnosis was recorded in 54.5% and more than two thirds had moderate to severe cognitive impairment (68.8%). Sixty-one percent had received at least one course of antibiotics in the 12 months prior to enrolment.

**Findings to date:** To investigate the representation of the GRACE cohort to Australians in residential aged care, its characteristics were compared to a subset of the historical cohort of the Registry of Senior Australians (ROSA). This included 142,923 individuals who were permanent residents of RACFs on June 30th, 2017. GRACE and ROSA cohorts were similar in age, sex, and duration of residential care, prevalence of health conditions, and recorded dementia diagnoses. Differences were observed in care requirements and antibiotic exposure (both higher for GRACE participants). GRACE participants had fewer hospital visits compared to the ROSA cohort, and a smaller proportion were prescribed psycholeptic medications.

**Future plans:** Participant and built environment metagenomes will be used to determine microbiome and resistome characteristics. Individual and facility risk exposures will be aligned with metagenomic data to identify principal determinants for AMR carriage. Ultimately, this analysis will inform measures aimed at reducing the emergence and spread of antibiotic resistant pathogens in this high-risk population.

**Strengths and limitations of this study:**

- The GRACE study captured a diverse array of data; demographics, medications, personal and medical care, RACF management practices, as well as oropharyngeal, intestinal, and environmental metagenomic data, allowing detailed analysis of exposure-resistome relationships.
- A high rate of participant recruitment (75% of eligible residents) was achieved, representing the spectrum of resident characteristics and care needs. This included a representative proportion of individuals with moderate or severe cognitive impairment.
- The main limitation of this cohort resulted from the early cessation of recruitment, due to stringent facility access regulations resulting from the COVID-19 pandemic. While a high recruitment rate partially compensated in terms of cohort size, we were unable to complete recruitment at our fifth site or begin recruitment at two further sites.
- Ethnic and linguistic data was not captured and so could not be compared between cohorts.

## INTRODUCTION

In keeping with trends globally, Australia is experiencing significant ageing of its population.^1^ By 2031, 21% of Australians will be aged over 65 years.^2^ Of Australians over 65, 6% currently live in residential aged care facilities (RACFs), and of those 85 years and over, 30% do.^2 3^ The threat of increasing rates of infection caused by multidrug-resistant organisms (MDRO) is particularly serious in RACFs. High rates of antibiotic prescription, poor antimicrobial stewardship, and the potential for microbial transmission between residents, all contribute to growing rates of multidrug-resistant clinical isolates.^4-6^ However, the prevalence of antimicrobial resistance (AMR) in asymptomatic individuals (carried either by pathogens or commensal microbes), or the dispersal of MDRO within the RACF environment, is largely uncharacterised. Despite serious concerns about a growing inability to readily treat common infections, and the potential for RACF populations to contribute to AMR carriage within the wider community, sufficiently detailed data to support the development of effective measures to limit the spread of MDRO in aged care simply do not exist.

The Generating evidence on Resistant bacteria in the Aged Care Environment (GRACE) study enrolled residents from five RACF located in metropolitan Adelaide, South Australia and aims to address five questions that are fundamental to developing strategies to reduce AMR carriage in RACF residents: 1) What factors determine the types and levels of AMR determinants carried by RACF residents? 2) To what extent is there evidence of AMR transmission between RACF residents? 3) Is interaction with the RACF built environment likely to facilitate AMR transmission? 4) Do hospital visits for acute care significantly influence types and levels of AMR carriage? 5) To what extent do ageing-associated changes in gut microbiology influence AMR carriage?

To address these research questions, participants were invited to provide stool and oropharyngeal samples for metagenomic analysis to determine microbiome and resistome characteristics. Environmental samples were also collected from areas within each facility. Metagenomic data will be related to a range of factors, including facility variables, resident demographics, morbidity, and polypharmacy data, to identify influences on AMR carriage and potential transmission.

Prior to these analyses, we compared GRACE cohort characteristics with those of aged care residents within the national historical cohort of the Registry of Senior Australians (ROSA) which contains data for more than 2.8 million Australians aged over 65 who accessed government-subsidised aged services from 1997 to 2017.^7^ Our comparison assessed whether the GRACE cohort was representative of the wider Australian aged care population, and its validity as a basis to provide further insight.

## COHORT DESCRIPTION

### Study design and population

GRACE is a prospective, cohort study of permanent residents of RACFs recruited between March 2019 and March 2020. All eligible residents living in participating facilities at the time of recruitment and/or their next of kin were approached by a research nurse to provide informed consent. In addition to the consent form, study information was made available in the form of a video, a two-page brochure and on a website. Consent could be provided for one or all study procedures, including the collection of stool and/or oropharyngeal samples, collection of facility-level medical records and access to data held by the Medicare Benefits Schedule (MBS) and Pharmaceutical Benefits Scheme (PBS). Participants were not eligible if: 1) they were in respite care, 2) they were receiving palliative/end-of-life care, 3) it was recommended by management that they not be approached, and 4) we were unable to contact next of kin where third-party consent was required. Participants who required third-party consent, such as those with cognitive impairment, were identified by the participating facility and communicated to the study team. GRACE aimed to recruit 400 residents across 10 RACFs. However, due to the COVID-19 pandemic, and the imposition of strict facility entry restrictions, recruitment was ceased, resulting in a sample size of 279 residents from five facilities, with a mean recruitment rate of 75%. Site 1 was excluded from this mean as data on eligibility and participants who declined was not recorded.

Of 403 residents assessed for eligibility, 344 were approached to join the study and 279 consented to participate (Figure 1). Fifty-nine residents were ineligible and 65 declined to participate (excluding site 1). Of those who consented, 111 (39.8%) provided consent themselves, and 168 (60.2%) provided third-party consent. Two-hundred and seventy-three residents (97.8%) provided consent for Department of Human Services (DHS) data access, with MBS and PBS data available for 243 and 228 residents, respectively.

**Figure 1.**
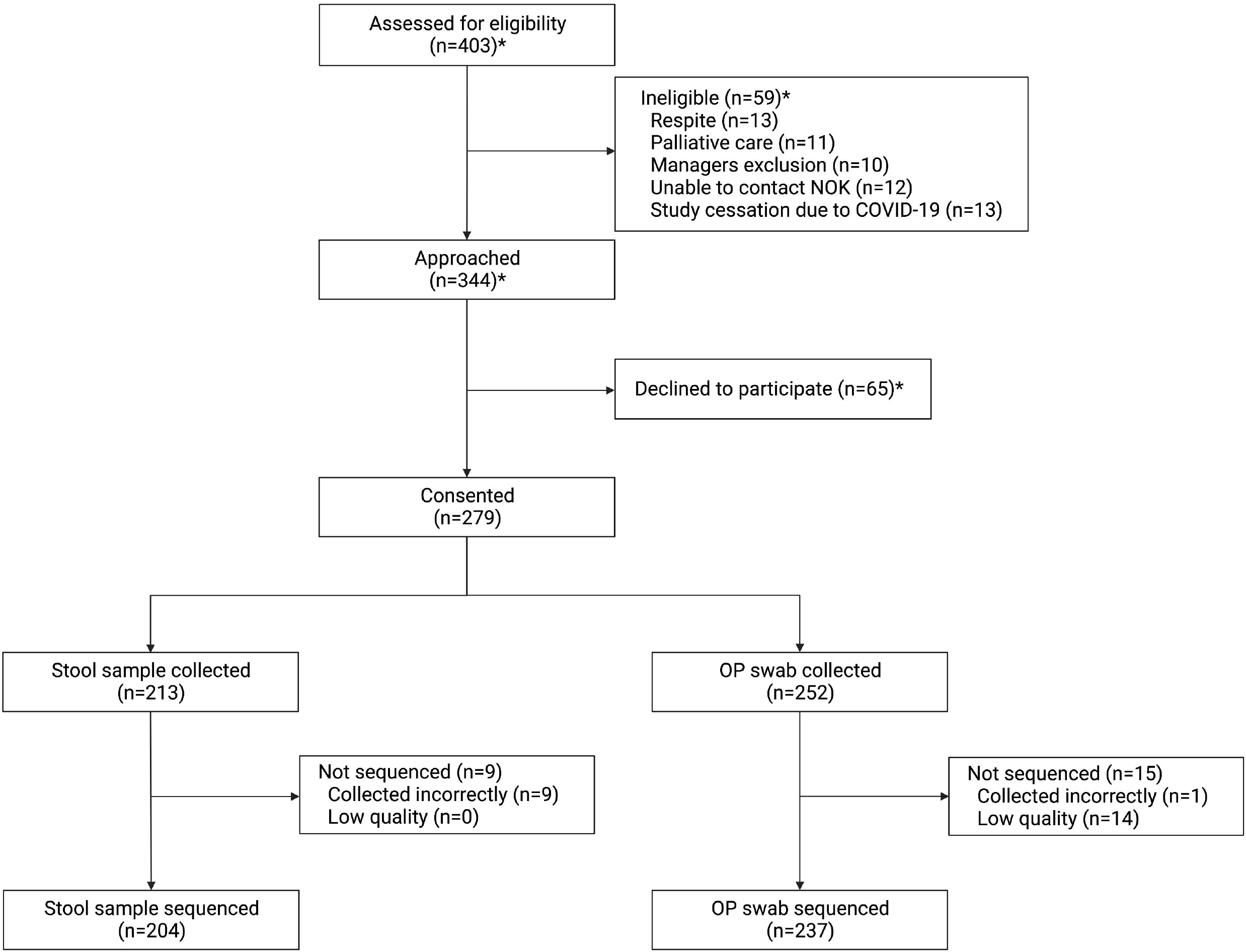
GRACE study recruitment and sample collection numbers. * data does not include site 1; NOK = next of kin; OP = oropharyngeal.

### Data collection

Participant and facility data were collected at the close of recruitment at each site and included facility medical records. Information held by the PBS and MBS from the DHS was requested after all recruitment was complete. Demographical data (including age and sex), as well as data on participant living arrangements (time spent in current facility, room type, room security) were collected from facilities. In addition, data on care requirements was collected from facilities via the Aged Care Funding Instrument (ACFI), a tool used on entry to an RACF to determine the funding needed for a person’s care.^8^ This includes three domains representing different areas of care needs: Activities of Daily Living, Cognition and Behaviour, and Complex Healthcare. Activities of Daily Living, includes details of care required for eating, showering, toileting and general mobility. Cognition and Behaviour domain measures the cognitive skills, verbal and physical behaviour, and mental health of individuals. Complex Healthcare considers the support residents need to manage their medications and health conditions. The ACFI also includes data on cognitive and behavioural conditions, which we have used to determine the presence of dementia in our cohort. Cognitive impairment scores pre-calculated using the Psychogeriatric Assessment Scales – Cognitive Impairment Scale (PAS-CIS) method were also obtained from the ACFI data.^9^ Details of hospitalisations in the 12 months prior to enrolment, diet type and texture, and medical care data (wound care, medical devices) were collected from the facility records.

Data collected from the PBS included medications prescribed during the 12 months prior to study enrolment for each participant. Specifically, data were obtained relating to medications that might directly or indirectly influence the microbiome and care needs (antibiotics, antivirals, antimycotics, medicines for constipation and acid-related disorders, insulin, antidiabetics, opioids, anti-inflammatories, corticosteroids, immunosuppressants, hormones, lipid-modifying and beta-blocking agents, antidementia medication, antidepressants, and psycholeptics (includes antipsychotics, anxiolytics, sedatives/hypnotics)). Data collected from the MBS included general practitioner (GP) attendances, specialist attendances, allied health services, surgery, diagnostic imaging services, health assessments, and access of pathology services during the 12 months prior to study enrolment for each participant. Definitions and coding of these variables can be found in Supplementary Table 1.

**Table 1.**
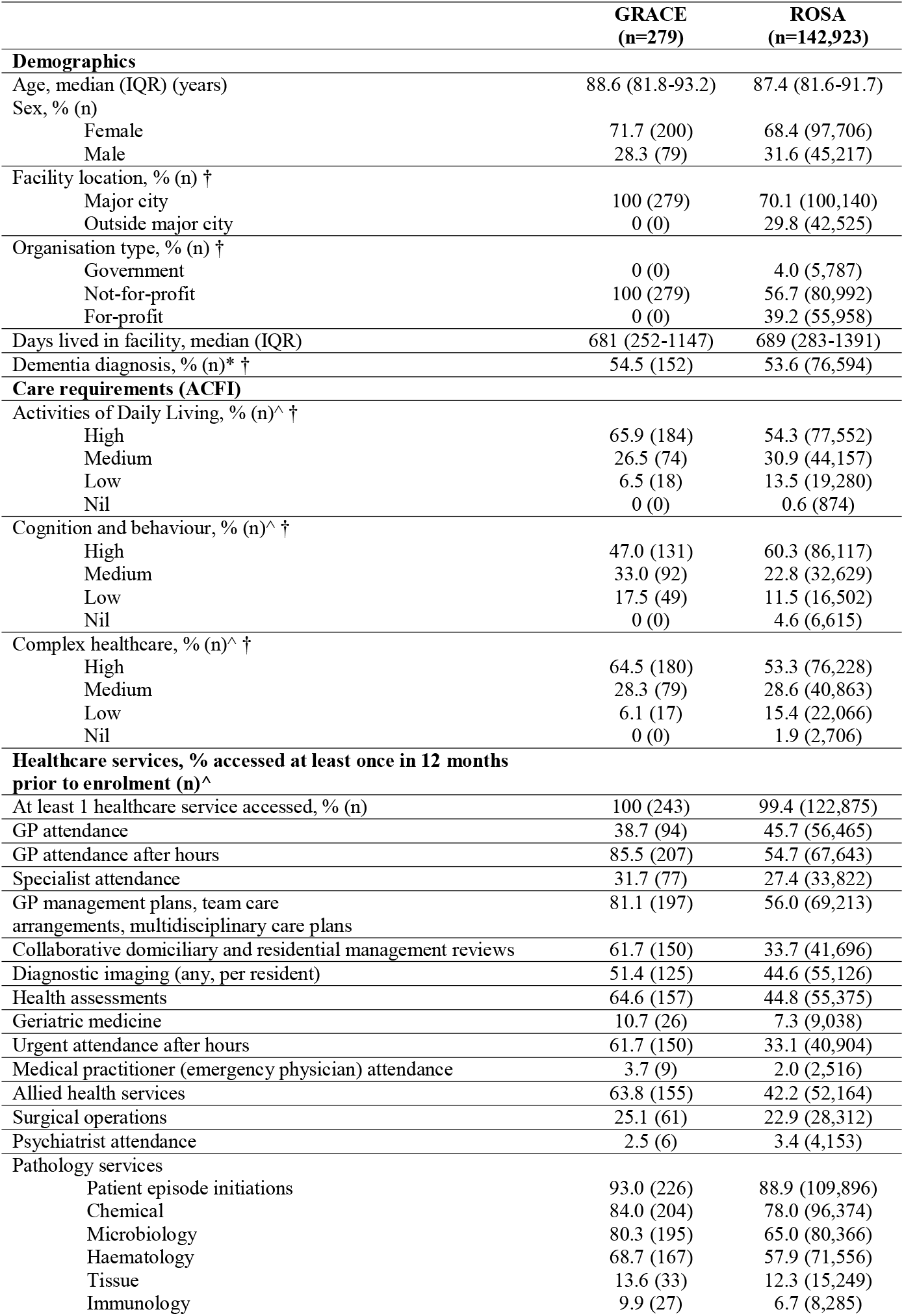
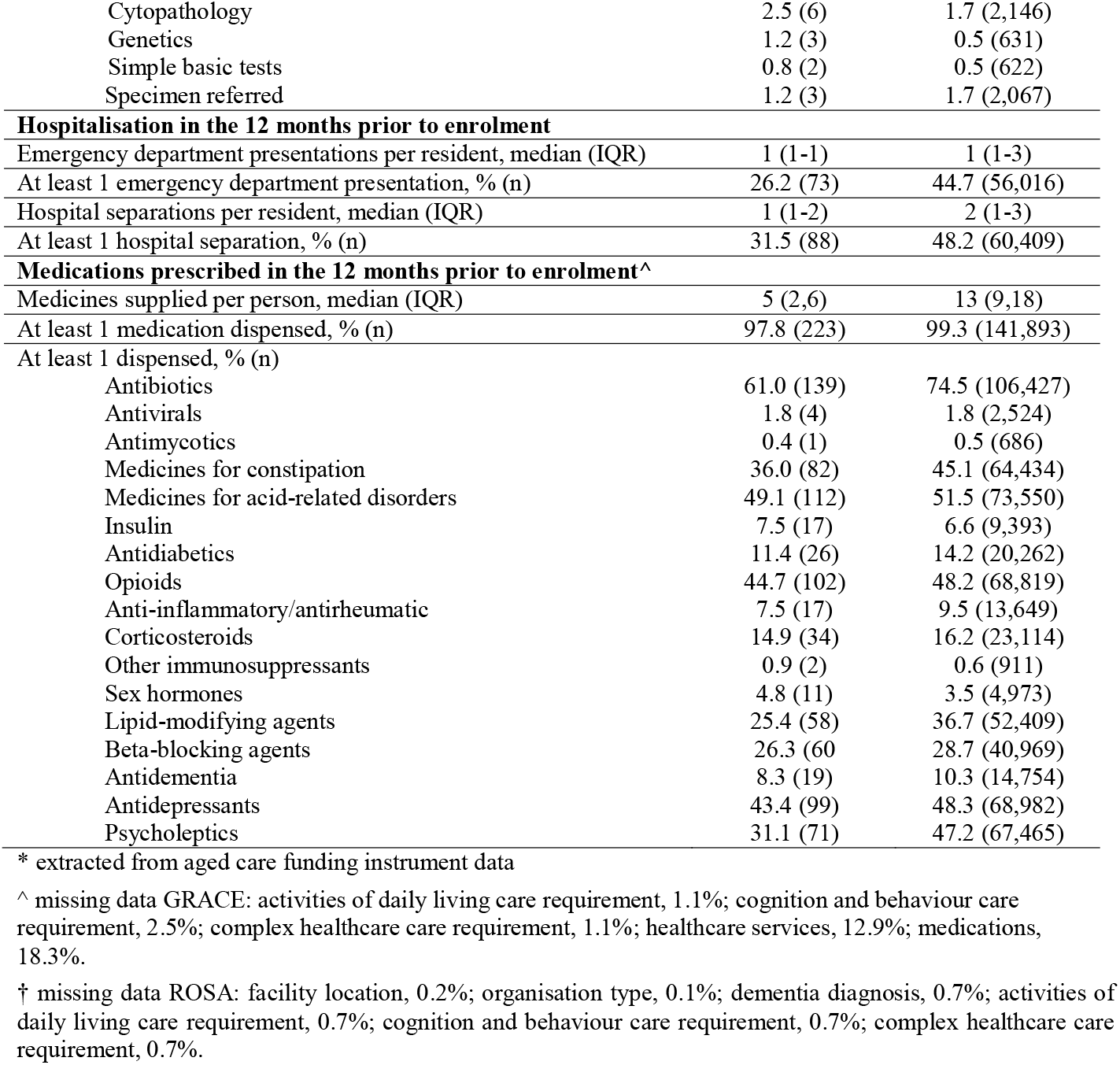
Characteristics of GRACE study participants compared to the national population in ROSA.

RxRisk is an established tool to determine a person’s actively managed health conditions using their medication data and was used to compare health conditions between GRACE and ROSA.^10^ In GRACE, RxRisk health conditions were able to be assessed for 228 participants as this relied on PBS data availability. Dementia is reported using both the RxRisk method and the ACFI diagnosis as per previously reported.^7^

### Comparison with the national aged care data

Data from the National Historical Cohort of the Registry of Senior Australians (ROSA) was used to evaluate the extent to which the study cohort was representative of the national residential aged care population.^7^ ROSA includes Australians aged 65 years and over who accessed government-subsidised aged care services between 1997 and 2017. ROSA has integrated information from the aged care sector with various health care data sources. Datasets within ROSA include: Australian Institute of Health and Welfare’s National Aged Care Data Clearinghouse datasets, Australian Government Medicare Benefits Schedule (MBS) and Pharmaceutical Benefits Scheme (PBS), state health authorities’ hospitalisations (QLD, NSW, VIC, SA), and ambulance datasets (NSW, SA). All data were de-identified and integrated by approved agencies (Australian Institute of Health and Welfare, Centre for Health Record Linkage, Centre for Victorian Data Linkage, SA NT DataLink and Queensland Health’s Statistical Services Branch). Details of ROSA datasets, variables, definitions, and limitations have been published previously.^7^ The June 30^th^, 2017 (latest available data at the time of the study), non-Indigenous national cohort of permanent residents of RACFs (n=142,923) was obtained from ROSA for comparison to the GRACE cohort. Analysis focusing on MBS subsidized health care services only included individuals without Department of Veterans’ Affairs cards (n=123,555), and analysis focusing on hospitalization records only included individuals living in NSW, VIC, SA, and QLD (n=125,351).

### Statistical Analysis

Descriptive statistics were used to summarise characteristics of both GRACE and ROSA derived populations. For continuous data, median (IQR) was reported. For categorical data, percent and number of participants was reported. GRACE data was exported, cleaned, and analysed in Statistical Analysis Software (SAS) University Edition (SAS Studio v3.8/SAS v9.4).

## FINDINGS TO DATE

### Demographics

A comparison of participant clinical data between GRACE (n=279) and ROSA (n=142,923) is shown in Table 1. GRACE participants had a median age of 88.6 (IQR=81.8-93.2) years, which was similar to that of ROSA (med=87.4, IQR=81.6-91.7). GRACE and ROSA participants were mostly female (GRACE=71.7%, n=200; ROSA=68.4%, n=97,706), had a similar prevalence of dementia (GRACE=54.5%, n=152, ROSA=53.6%, n=76,594), and residents had been in their current facility for a similar period of time at recruitment/data collection (GRACE: med=681 days, IQR=252-1147; ROSA: med=689 days, IQR=283-1391). GRACE participants all lived in metropolitan facilities, run by not-for-profit organisations, whereas ROSA participants lived in a number of locations and organisation types.

### Care requirements

Care requirements represented by the three ACFI domains were assessed for both datasets (Table 1). Activities of Daily Living (ADL) care requirements were greater in the GRACE cohort compared to ROSA, with 65.9% (n=184) having a high care requirement for this domain, compared to 54.3% in ROSA (n=77,552). Cognition and Behaviour care requirements for GRACE were less than those in ROSA, with 47.0% (n=131) and 60.3% (n=86,117) having a high care score, respectively. GRACE had a higher proportion of participants with a high care requirement for Complex Healthcare (64.5%, n=180) compared to the ROSA cohort (53.3%, n=76,228).

### Utilisation of healthcare services subsidised by Medicare

In the 12 months prior to enrolment/data collection, the proportions of the GRACE and ROSA cohorts that had accessed an MBS-subsidised healthcare service were similar (GRACE=100%, n=243; ROSA=99.4%, n=122,875). GRACE participants utilised GP services for non-urgent out of hours care most commonly (GRACE=85.5%, n=207; ROSA=54.7%, n=67,643; Table 1). More GRACE participants accessed urgent out of hours GP services (61.7%, n=150) compared to ROSA (33.1%, n=40,904). Both cohorts accessed standard GP attendances similarly (GRACE=38.7%, n=94; ROSA=45.7%, n=56,465). GRACE participants had team care plans (in which multidisciplinary teams manage a case; GRACE=81.1%, n=197; ROSA=56.0%, n=69,213) and collaborative domiciliary and residential management reviews (in which a GP and pharmacist review ongoing medication for a resident; GRACE=61.7%, n=150; ROSA=33.7%, n=41,696) more commonly than those in the ROSA.

GRACE and ROSA cohorts had a similar level of pathology service utilisation, with patient episode initiations the most frequently accessed service for each (GRACE=93.0%, n=226; ROSA=88.9%, n=109,896; Table 1). Of all pathology services captured, access of microbiology services differed most between the datasets (GRACE=80.3%, n=195; ROSA=65.0%, n=80,366).

### Hospitalisations

GRACE had a smaller proportion of participants with at least 1 hospitalisation recorded in the 12 months prior to enrolment/data collection (31.5%, n=88) compared to ROSA (48.2%, n=60,409) The median number of hospitalisations per resident was similar (GRACE: med=1, IQR=1-2; ROSA: med=2, IQR=1-3; Table 1). GRACE also had a smaller proportion of participants with at least 1 emergency department presentation in the 12 months prior to enrolment/data collection (GRACE=26.2%, n=73; ROSA=44.7%, n=56,016).

### Medications

At least 1 medication had been dispensed in the 12 months prior to enrolment/data collection for 97.8% (n=223) and 99.3% (n=141,893) of the GRACE and ROSA cohorts, respectively. GRACE participants were taking less medications per person (med=5, IQR=2-6) compared to ROSA (med=13, IQR=9-18; Table 1). Antibiotics were the most commonly supplied drug class to both cohorts during the 12 months prior to enrolment/data collection, but GRACE had a fewer number of participants who were supplied antibiotics (61.0%, n=139) compared to ROSA (74.5%, n=106,427). Psycholeptics were supplied to fewer participants in GRACE (31.1%, n=71) compared to ROSA (47.2%, n=67,465).

### Health conditions

The median number of RxRisk conditions per participant did not differ between the two cohorts (both: med=5, IQR=3-7; Table 2). In GRACE, the most common RxRisk conditions included gastro-oesophageal reflux disease (49.1%, n=112), pain (44.7%, n=102) and depression (43.0%, n=98). Gastro-oesophageal reflux disease was also the most common condition in ROSA (49.0%, n=69,977), followed by depression (45.7%, n=65,351) and hypertension (43.1%, n=61,542). Compared to ROSA, the GRACE cohort had a higher proportion of participants being treated for osteoporosis/Paget’s disease (GRACE=23.7%, n=54; ROSA=15.6%, n=22,306) and hypothyroidism (GRACE=17.5%, n=40; ROSA=10.8%, n=15,450). Most conditions were similar in their prevalence between the two datasets.

**Table 2.** RxRisk-V health conditions for GRACE participants compared to ROSA.

|  | **GRACE**<br>**(n=228)** | **ROSA**<br>**(n=142,923)** |
| --- | --- | --- |
| **RxRisk-V condition, % (n)^** |  |  |
| Gastro-oesophageal reflux disease | 49.1 (112) | 49.0 (69,977) |
| Hyperlipidaemia | 25.4 (58) | 32.1 (45,927) |
| Hypertension | 40.4 (92) | 43.1 (61,542) |
| Ischaemic heart disease: hypertension | 32.9 (75) | 35.6 (50,863) |
| Antiplatelets | 19.3 (44) | 19.1 (27,258) |
| Depression | 43.0 (98) | 45.7 (65,351) |
| Pain | 44.7 (102) | 41.2 (58,921) |
| Anticoagulants | 21.9 (50) | 17.4 (24,838) |
| Chronic airways disease | 24.1 (55) | 22.0 (31,493) |
| Congestive heart failure | 18.4 (42) | 17.0 (24,362) |
| Osteoporosis/Paget | 23.7 (54) | 15.6 (22,306) |
| Psychotic illness | 13.2 (30) | 15.9 (22,680) |
| Diabetes | 15.0 (34) | 15.4 (22,025) |
| Steroid responsive disease | 14.0 (32) | 10.2 (14,525) |
| Arrhythmia | 10.5 (24) | 10.1 (14,389) |
| Anxiety | 17.1 (39) | 14.5 (20,742) |
| Glaucoma | 11.0 (25) | 11.3 (16,120) |
| Ischaemic heart disease: angina | 11.0 (25) | 9.7 (13,812) |
| Dementia | 8.3 (19) | 16.5 (23,520) |
| Hypothyroidism | 17.5 (40) | 10.8 (15,450) |
| Inflammation/pain | 7.5 (17) | 5.8 (8,247) |
| Gout | 7.0 (16) | 5.9 (8,403) |
| Parkinson's disease | 11.8 (27) | 7.3 (10,435) |
| Epilepsy | 7.0 (16) | 9.2 (13,167) |
| Liver failure | 0 (0) | 0.1 (81) |
| Incontinence | 3.5 (8) | 2.8 (4,063) |
| Benign prostatic hyperplasia | 3.1 (7) | 2.8 (4,064) |
| Malignancies | 1.3 (3) | 1.5 (2,174) |
| Renal disease | 2.6 (6) | 1.1 (1,630) |
| Hyperthyroidism | 0 (0) | 0.9 (1,304) |
| Allergies | 0 (0) | 0.6 (842) |
| Migraine | 0 (0) | 0.6 (810) |
| Irritable bowel syndrome | 0.4 (1) | 0.6 (831) |
| Smoking cessation | 0.4 (1) | 0.4 (581) |
| Pancreatic insufficiency | 0 (0) | 0.4 (547) |
| Psoriasis | 1.3 (3) | 0.5 (655) |
| Bipolar disorder | 0 (0) | 0.5 (675) |
| Transplant | 0 (0) | 0.1 (145) |
| Alcohol dependency | 0 (0) | <0.1 (39) |
| Pulmonary hypertension | 0 (0) | <0.1 (37) |
| Hepatitis B | 0 (0) | <0.1 (19) |
| HIV | 0 (0) | <0.1 (38) |
| Hyperkalaemia | 0 (0) | <0.1 (20) |
| Malnutrition | 0 (0) | 0 (0) |
| Tuberculosis | 0 (0) | <0.1 (<5) |
| Hepatitis C | 0 (0) | <0.1 (10) |
| Number of conditions per person, median (IQR) | 5 (3,7) | 5 (3,7) |

^ missing data GRACE, 18.3%.

### Additional GRACE datapoints

Some characteristics could not be compared between the GRACE and ROSA cohorts and these are summarised in Supplementary Table 2. Most GRACE participants were staying in their own rooms (97.8%; n=273), with a small proportion also living in memory support areas (12.9%; n=36). Diet type and texture was highly conserved among participants, with 93.9% (n=262) reporting a normal diet, 72.8% reporting normal meal texture (n=203) and 91.4% (n=255) reporting a normal liquid texture. Most participants were receiving a standard fortified diet (56.3%; n=157), but a large proportion were receiving a high energy high protein supplemented diet (39.4%; n=110). Seven (2.5%) participants reported a colostomy/ileostomy. No GRACE participants reported a urinary catheter *in situ*, vascular catheter *in situ*, urostomy, or a tracheostomy hence they are not included in Supplementary Table 2. Seventy-two (26.0%) participants were receiving wound care at the time of enrolment, with grade 1-2 pressure ulcers the most common wound being treated (6.5%; n=18). Using the pre-calculated PAS-CIS method, GRACE participants were most commonly assigned a moderate impairment score (39.8%; n=111), followed by severe (28.0%; n=78) and mild impairment (27.6%, n=77). Very few were assigned no to minimal impairment (2.9%; n=8).

## STRENGTHS AND LIMITATIONS

The primary strength of the GRACE study is the combination of comprehensive demographic, health care, health status, medical, pharmaceutical, and facility variables with intestinal and oropharyngeal microbiome and resistome data for permanent residents of RACFs. The research design developed to establish this cohort has enabled powerful opportunities for novel and extensive investigations currently underway into relationships between risk factors in aged care, current practices in aged care, intestinal health, and disease state outcomes. For example, future work will include investigations assessing associations between cognitive and behavioural diagnoses and the composition of the microbiota to measure the impact of variables in aged care on the increasing burden of cognitive decline.

Another key strength from GRACE was the high rate of recruitment (75%) from residents and families of residents in RACFs. This was most likely attributable to the availability of a research nurse in the study team who personally and extensively communicated with residents and their families. For enhanced rates of recruitment, future studies may also benefit from dedicating significant resources towards communication strategies, particularly when involving elderly populations.

A limitation of the GRACE study resulted from the impact of the SARS-CoV-2 (COVID-19) pandemic. However, whilst this affected the final sample size, the severity of the effect was dampened by the high recruitment rate of participants. In addition, the location of participating RACFs should be considered. These were exclusively in metropolitan areas and may therefore differ in their characteristics from those located in rural or more remote areas. Similarly, only not-for-profit aged care providers participated in the study, who may have had higher staffing capabilities and different approaches to food provisions compared to other types of facilities. Subsequent studies would benefit from a diversified cross-section of

RACFs in both geographical location and funding type. Data relating to resident ethnicity was also not captured.

Specifically in relation to the emergence of antibiotic resistance, potential risk exposures, including high antibiotic use, high medication usage, a high proportion of health conditions experienced, and frequent access of after-hours GP services, were identified in the GRACE cohort, and were reflective of exposures in the wider residential aged care population.

## Collaboration

The GRACE team have established a cohort with comprehensively detailed information on overall health, microbiome profiles, and medication use in RACFs. The primary aim for establishing this cohort was to investigate the existence and spread of resistant bacteria in residential aged care to help improve facility management, prevent the spread of harmful bacteria, and ultimately improve the health of aged care residents and the wider community. The authors welcome approaches from other researchers to discuss the potential for collaborative studies that utilise this valuable resource.

## Data Availability

The GRACE study data are available upon reasonable request. GRACE study data described in this article are available to all interested researchers through collaboration. Please contact GR. Metagenomic sequencing data will be made available via a public registry once completed. Due to data custodian restrictions related to the sharing of linked data in ROSA, these data cannot be made publicly available to other researchers.

## Abbreviations

GRACE: Generating evidence on antimicrobial Resistance in the Aged Care Environment
ROSA: Registry Of Senior Australians
RACF: Residential Aged Care Facility
AMR: Antimicrobial Resistance
MDRO: Multi-Drug Resistant Organism
PBS: Pharmaceutical Benefits Scheme
MBS: Medicare Benefits Schedule
DHS: Department of Human Services
ACFI: Aged Care Funding Instrument
PAS-CIS: Psychogeriatric Assessment Scales – Cognitive Impairment Scale
GP: General Practitioner

## Acknowledgements

The authors are grateful to the participants and their families, and staff of the residential aged care facilities who participated in the GRACE study. We would like to acknowledge Registry of Senior Australians’ (ROSA) Steering Committee and the ROSA South Australian Health and Medical Research Institute (SAHMRI) Research Team for ensuring the success of the ROSA and support with this study. We also acknowledge the South Australian Government Department for Innovation and Skills (2017-2021) who provided support to establish ROSA, the Australian Government Medical Research Future Fund (2021-2024, PHRDI000009), and ROSA collaborating partners (SAHMRI, ECH Inc, Silver Chain, Life Care) for its ongoing support, and the Australian Institute of Health and Welfare for the linkage and construction of input data, SA Health, NSW Ministry of Health, Victorian Department of Health and QLD Health for the provision of the state-based data used in the ROSA with linkage via the AIHW, Centre for Health Record Linkage (CHeReL), the Centre for Victorian Data Linkage (CVDL), SA NT DataLink and Queensland Health’s Statistical Services Branch.

## Contributors

LC, EF, LP, MC, MI, and GR conceived and designed the study. LC, CL, EF, and JC acquired the data. LC and CL analysed and interpreted the data. LC, AS and EF drafted the manuscript. All authors provided intellectual input to the manuscript and critically revised the manuscript. All authors have read and approved the final version of the manuscript.

## Patient and public involvement

RACF staff provided advice on how to engage with participants, families, and other staff members, and the formulation of the participant information data capture instrument. Although residents were not involved in the design, recruitment, conduct, reporting, or dissemination of this study directly, resident committees were involved through presentations at their respective sites. Ongoing feedback is provided to aged care providers and their participating residents via printed, electronic, and in-person communication.

## Patient consent for publication

Not applicable.

## Ethics approval

Ethics approval for the GRACE study was obtained from the Southern Adelaide Clinical Human Research Ethics Committee (HREC/18/SAC/244). Participants provided written informed consent themselves or where third-party consent was required, a legal guardian or family member with power of attorney provided consent on their behalf. ROSA has been reviewed and approved by the following Human Research Ethics Committees (HREC): University of South Australia HREC (Ref: 200487; October 2017), Australian Institute of Health and Welfare (AIHW Ref: EO2018/1/418; February 2018), the SA Department for Health & Wellbeing HREC (Ref: HREC/18/SAH/90; November 2018), the NSW Population and Health Services Research Ethics Committee (Ref: 2019/ETH12028; September 2019) and the Aboriginal Health Research Ethics Committee (AHREC Ref 04-20-895; September 2020).

## Provenance and peer review

Not commissioned.

