## Supplemental Information for "Cohort Profile: The Australian Generating evidence on antimicrobial resistance in the aged care environment (GRACE) study; alignment with national population characteristics"

**SUPPLEMENTARY MATERIALS**

**Supplementary Table 1.** Health services, pathology, and medicines coding.

| **Description** | **Code** |
| --- | --- |
| **Health Services** | **MBS group code** |
| GP attendance to which no other item applies | A01 |
| GP after hours attendances to which no other item applies | A22 |
| Specialist attendance | A03 |
| GP management plans, team care arrangements, multidisciplinary care plans | A15 |
| Collaborative domiciliary and residential management reviews | A17 |
| Diagnostic imaging^ | I* |
| Health assessments | A14 |
| Geriatric medicine | A28 |
| Urgent attendance after hours | A11 |
| Medical practitioner (emergency physician) attendance to which no other item applies | A21 |
| Allied health services | M03 |
| Surgical operations | T08 |
| Psychiatrist attendance | A08 |
| **Pathology Services** | **MBS group code** |
| Patient episode initiations | P10 |
| Chemical | P02 |
| Microbiology | P03 |
| Haematology | P01 |
| Tissue | P05 |
| Immunology | P04 |
| Cytopathology | P06 |
| Genetics | P07 |
| Simple basic tests | P09 |
| Specimen referred | P11 |
| **Medicines** | **ATC code** |
| Antibiotics | J01* |
| Antivirals | J05* |
| Antimycotics | J02* |
| Medicines for constipation | A06* |
| Medicines for acid-related disorders | A02* |
| Insulin | A10A* |
| Antidiabetics | A10B* |
| Opioids | N02A* |
| Anti-inflammatory/antirheumatic | M01* |
| Corticosteroids | H02* |
| Other immunosuppressants | L04* |
| Sex hormones | G03* |
| Lipid-modifying agents | C10* |
| Beta-blocking agents | C07* |
| Antidementia | N06D* |
| Antidepressants | N06A* |
| Psycholeptics | N05* |

^Any MBS group in the diagnostic imaging category

**Supplementary Table 2**. Additional characteristics of GRACE participants.

|  | **Total**  **(n=279)** |
| --- | --- |
| **Room detail** |  |
| Room type, % (n) |  |
| Single | 97.8 (273) |
| Shared | 2.2 (6) |
| Memory support room, % (n) |  |
| Yes | 12.9 (36) |
| No | 87.1 (243) |
| **Diet** |  |
| Diet type, % (n)^ |  |
| Normal | 93.9 (262) |
| Vegetarian | 0.4 (1) |
| Lactose free | 3.9 (11) |
| Gluten free | 0.7 (2) |
| Lactose and gluten free | 0.7 (2) |
| Meal texture, % (n) |  |
| Regular | 72.8 (203) |
| Finger food | 0.4 (1) |
| Soft | 12.9 (36) |
| Minced and moist | 7.9 (22) |
| Pureed | 6.1 (17) |
| Liquidised | 0 (0) |
| Liquid texture, % (n) |  |
| Normal/Thin | 91.4 (255) |
| Slightly thick | 1.4 (4) |
| Mildly thick | 5.0 (14) |
| Moderately thick | 1.4 (4) |
| Extremely thick | 0.7 (2) |
| Prescribed supplementation, % (n)^ |  |
| Standard fortified diet | 56.3 (157) |
| High energy & high protein | 39.4 (110) |
| Oral supplement | 0 (0) |
| PEG supplement | 0 (0) |
| Multiple | 1.1 (3) |
| **Medical Care** |  |
| Colostomy/ileostomy, % (n) |  |
| Yes | 2.5 (7) |
| No | 97.5 (272) |
| Wound care, % (n)^ |  |
| Not receiving wound care | 73 (205) |
| Receiving care for multiple wounds | 3.2 (9) |
| Skin tear | 5.4 (15) |
| Pressure ulcer (grade 1-2) | 6.5 (18) |
| Pressure ulcer (grade 3-4) | 0.7 (2) |
| Leg ulcer | 1.8 (5) |
| Burn/scald | 0 (0) |
| Abrasion/graze | 2.2 (6) |
| Surgical wound | 0.7 (2) |
| Lesion | 0.7 (2) |
| Unspecified | 4.7 (13) |
| **Cognitive Impairment** |  |
| Cognitive impairment level, % (n)^ |  |
| None/Minimal | 2.9 (8) |
| Mild | 27.6 (77) |
| Moderate | 39.8 (111) |
| Severe | 28.0 (78) |

**^** missing data GRACE: diet type, 0.4%; prescribed nutritional supplement, 3.2%; receiving wound care, 0.7%; cognitive impairment level, 1.8%.
